# A Dynamical Map to Describe COVID-19 Epidemics

**DOI:** 10.1101/2021.03.10.21253322

**Authors:** Eduardo V. M. dos Reis, Marcelo A. Savi

## Abstract

Nonlinear dynamics perspective is an interesting approach to describe COVID-19 epidemics, providing information to support strategic decisions. This paper proposes a dynamical map to describe COVID-19 epidemics based on the classical susceptible-exposed-infected-recovered (SEIR) differential model, incorporating vaccinated population. On this basis, the novel map represents COVID-19 discrete-time dynamics by adopting three populations: infected, cumulative infected and vaccinated. The map promotes a dynamical description based on algebraic equations with a reduced number of variables and, due to its simplicity, it is easier to perform parameter adjustments. In addition, the map description allows analytical calculations of useful information to evaluate the epidemic scenario, being important to support strategic decisions. In this regard, it should be pointed out the estimation of the number death cases, infectious rate and the herd immunization point. Numerical simulations show the model capability to describe COVID-19 dynamics, capturing the main features of the epidemic evolution. Reported data from Germany, Italy and Brazil are of concern showing the map ability to describe different scenario patterns that include multi-wave and plateaus behaviors. The effect of vaccination is analyzed considering different campaign strategies, showing its importance to control the epidemics.

## 1. Introduction

The novel coronavirus disease (COVID-19) is a devastating pandemic with unprecedent consequences that are promoting a huge crisis all over the world. Due to that, scientific community is highly engaged, investigating the subject from different perspectives in distinct areas of human knowledge. COVID-19 dynamics shows a perspective that has an increasing interest due the possibility to understand pandemic evolution and to establish proper health strategy plans. Besides several publications, some journals are dedicating special issues putting togehter some of these efforts [1, 2].

Dynamical perspective is an interesting approach to deal with biomedical systems [3]. The literature presents several approaches regarding dynamical epidemic models [4], generally employed to describe different infectious diseases. An interesting and useful approach is based on population dynamics, where different populations are employed to represent the disease, establishing their evolution and interactions. Kermack and McKendrick [5] were one of the pioneers in considering three populations in epidemic models: susceptible, infected and recovered (SIR). This model describes the course of an epidemic showing that it cannot be terminated necessarily by all individuals becoming infected. The evolution of this approach was due to Anderson [6] and May [7] that considered an extra population: exposed (E). Nowadays, a well-established epidemic model is based on the susceptible-exposed-infected-removed (SEIR) framework, broadly employed to describe different infectious diseases as Influenza, Zika, HIV, among others. Additionally, a similar framework can be employed to describe other biological dynamics, such as the HTLV-I cell infection [8]

Hethcote [9] discussed the interpretation of the SEIR and other epidemic models. Zhao et al. [10] used an adapted version of SEIR model to include age groups, investigating the role of age on tuberculosis transmission. Dantas et al. [11] used SEIR-SEI model to describe the 2016 outbreak of Zika virus in Brazil, reporting the virus dynamics among the populations of humans and vectors. Some authors have also studied the mathematical aspects of the model, such as steady states and global stability [12, 13, 14]. Nowadays, these models are largely employed to describe COVID-19.

Specifically associated with COVID-19, Lin et al. [15] proposed a conceptual model for COVID-19 in Wuhan - China considering individual behavior reaction to the outbreak scenario and governmenatal actions. Ramos et al. [16] developed a novel mathematical model taking into consideration undeteced cases and different sanitary and infectiousness conditions of hospitalized people. This model was employed to study the evolution of COVID-19 in China and showed a good agreement with reported data. Savi et al. [17] applied the SEIR descritpion to investigate the pandemic evolution in Brazil, after a model verification with data from China, Italy and Iran. Results showed the importance of both governmental and individual actions to control the virus spread and to reduce the number of infected population. Pacheco et al. [18] improved the model including hospital infrastructure and explicitly spliting removed populations into recovered and deaths. Chen et al. [19] used an extended SIR model considering two types of infected persons: detectable and undetectable. China pandemic were treated showing that one-day prediction errors are almost less than 3%. Sujath [20] employed a machine learning method to forecast COVID-19 evolution in India employing linear regression, multilayer perception and vector autoregression methods.

From the mathematical point of view, SEIR epidemic models are governed by a system of ordinary differential equations, continuously describing the population evolution throughout time. An alternative to continuous models can be established by maps, that are discrete-time and governed by a system of algebraic equations. Dynamical maps have advantages due to their simplicity. Alonso-Quesada et al. [21] pointed out that the use of discrete-time instead of continuous-time models is preferred since the amount of necessary computation effort can be considerably reduced. Since epidemic statistics take place on fixed time intervals, it makes easier to parameterize a discrete-time than a continuous-time epidemic model. Enatsu et al. [22] stated that there are situations that constructing discrete epidemic models are more appropriate to understand disease transmission dynamics and to evaluate eradication policies since they permit arbitrary time-step units, preserving the basic features of corresponding continuous-time models.

Kwon and Jung [23] employed a discrete version of SEIR model to characterize the spread of coronavirus MERS in Korea, showing that an effective quarentine plan would reduce the maximum number of infected population by about 69% and MERS fade-out period may be shortened by about 30%. Din [24] analyzed the global stability analysis of the equilibrium points in the discrete-time form of SIR model. Enatsu et al. [22] used a backward differential scheme to discretize a class of SIR differential models showing that the effect of discretization is harmless to the global stability of the epidemic equilibrium. Alonso-Quesada et al. [21] proposed another method to discretize the SEIR method also considering natural births, deaths and reinfection. Cui et al. [25] built a discretized version of the SIR model using a nonstandard finite difference scheme. The model was applied to childhood diseases carrying out the vaccination effect of new borns, showing that the discrete and continuous models have the same equilibrium points. All these models are discrete versions of the continuous models.

The effect of vaccination is often considered in epidemic models in order describe the disease spread control. Different vaccination strategies can be imagined for each disease and their models consider that vaccination rate can be a function of either time or the susceptible population. Pulse vaccination is one kind of strategy characterized to be periodic in time [26, 27]. On the other hand, continuous vaccination strategy is the alternative that is descbribed by Gumel et al. [28] that showed the potential impact of SARS vaccine over the pandemic that spread to over 32 countries in 2003. Alexander et al. [29] built a model to study the transmission of influenza virus, computing the threshold vaccination rate necessary for community wide control. Kabir and Tanimoto [30] considered the effects of information buzz and information costs on the vaccination effect. Other references also discussed the effects of this kind of vaccination [31, 32].

This work deals with a novel COVID-19 dynamical map that describes the epidemics from infected, cumulative infected and vaccinated populations. The discrete-time model is developed based on the SEIRV model that employs differential equations to deal with the evolution of susceptible-exposed-infected-removed-vaccinated populations. The novel map reduces the six coupled ordinary differential equations into three algebraic equations, being capable to capture the main features of the COVID-19 epidemics with less model variables and parameters. In addition, the simplicity of the model allows analytical calculations of useful information to evaluate epidemic scenarios. In this regard, for instance, one should mention the infectious ratio and the herd immunization point, which are crucial information for strategic decision. The paper exploits the novel map through different perspectives. A model verification is carried out comparing its predictions with the classical SEIR differential model and reported data. In this regard, reported data from Germany, Italy and Brazil are analyzed showing good agreement between them and simulated results. A stability analysis is performed showing the conditions to control the epidemic spread and defining proper parameters for this aim. Different scenarios are investigated showing the rich dynamical perspective of COVID-19 epidemics that include multi-wave pattern with bell shape and plateaus characteristics. The effect of vaccination is carried out showing its importance to reduce the number of deaths considering different vaccination strategies. Results show that nonlinear dynamics perspective is an efficient tool to analyze COVID-19 epidemic evolution as long as a proper time scale is of concern.

## 2. Mathematical Model

This work has the main goal to develop a dynamical map to describe COVID-19 epidemics based on the classical SEIR framework. An extra population is incorporated considering the effect of vaccination, defining an SEIRV model, which considers the following populations: susceptible, *S*; exposed, *E*; active infected, *I*; removed, *R*, that accounts for recovered and deaths; and vaccinated, *V*. In addition, the cumulative infected population *C* is incorporated as a useful model information. An essential assumption is that infection can occur only once, which means that reinfection is neglected and, once vaccinated, an individual cannot become infected anymore. Although it is not possible to assure that this is a correct hypothesis for the COVID-19 epidemic, it can be considered reasonable valid for a time scale where the loss of immunity is long when compared with the period of analysis [16, 17]. In addition, it is assumed that the population *V* accounts only for the vaccinated population, which excludes situations where vaccination occurs after the infection. On this basis, the governing equations are defined as follows.

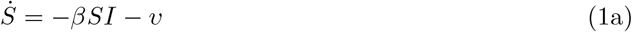

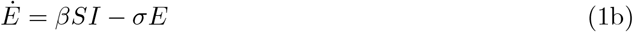

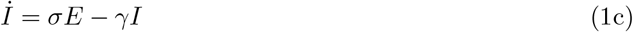

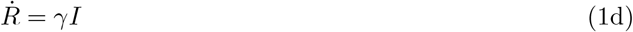

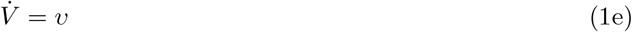

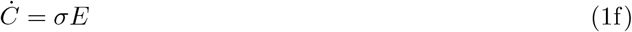

where dot represents time derivative; *β* is the transmission rate that is directly associated with social isolation; *σ*^*−*1^ is the mean latent period; *γ*^*−*1^ is the infectious period; and *υ* = *υ*(*S*) is the vaccination rate that is considered as a function of the susceptible population. It should be pointed out that dimensionless variables are considered and, therefore, (*S, E, I, R, V*) ∈ [0, 1] and *S* + *E* + *I* + *R* + *V* = 1. Since reinfection is neglected and since the average immunity period after vaccination is usually longer than the mean latent period, an exposed individual eventually becomes infected before acquiring immunity. Therefore, the vaccinated population comes from the interaction with susceptible population, *S*. The total cummulative infected population is associated with infected population since the change rate of both populations are due to the term *σE* [11, 17]. The map is derived from the SEIRV model represented by these differential equations, adopting some basic assumptions. By considering each one of the populations, represented by *X*, its time derivative is: 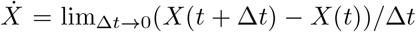, which means that 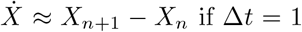 and *n* representing the *n*-th day. Under this assumption, the following steps are followed to build the dynamical map:

1. Substituting Eq. (1b) into Eq. (1c), one obtains: *İ* = *βSI* − *Ė* − *γI*.
2. It is assumed that the ratio *E/I* assumed in the beginning of the outbreak is kept constant throughout the whole epidemic period, which means that *E* = Λ*I* and *Ė* = Λ*İ*. Substituting both Eq. (1b) and Eq. (1c) into it, making *S* → 1 (onset of outbreak), it yields 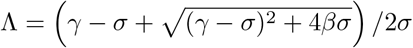.
3. Integrating Eq. (1d), 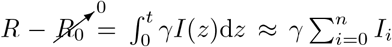, together with step 2 assumption (*E/I* = *Ė /İ* = Λ) into *S* + *E* + *I* + *R* + *V* = 1, one can write the susceptible group *S* as a function of *I*: *S* = 1 −(1 +Λ)*I* − *γ* Σ*i I*_*i*_ − *V*. Note that it is assumed *R*_0_ = 0 since *R*_0_ stands for the recovered group during the beginning of the outbreak.
4. Substituting Eq. (1f) into Eq. (1c), one obtains *Ċ* = *İ* + *γI*. Therefore, *C*_*n*+1_ = *C*_*n*_ + *I*_*n*+1_ + (*γ* − 1)*I*_*n*_.
5. By considering a generic *n*, summing all the time steps, one can find that 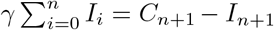, where it is assumed that *I*_0_ ≈ *C*_0_, since the summation must consider all the active cases from the beginning of the outbreak.
6. Since *S* + *E* + *I* + *R* = 1, the vaccination rate *υ*(*S*) can be expressed as *υ*(*I, C, V*).

After this sequence, it is possible to isolate both *I*_*n*+1_ and *C*_*n*+1_ to present the COVID-19 map as follows

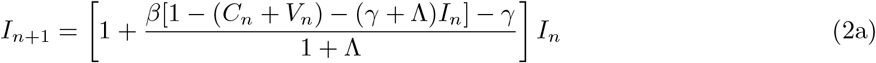

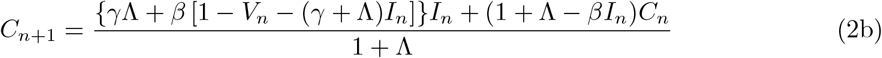

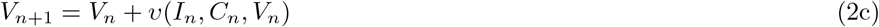

With

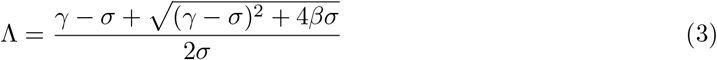

where Λ = *E/I* is a constant estimated by a parametric condition (*β, σ, γ*). It is important to highlight that, the population *E* was explicitly eliminated by assuming the ratio *E/I* constant, but its effect is implicitly present on the map dynamics. Moreover, despite this novel map is employed herein to describe COVID-19 dynamics, it can also be employed to describe the dynamics of any other epidemics.

The general form of the vaccination rate is a function of the infected, cumulative and vaccinated populations, *υ* = *υ*(*S*) = *υ*(*I, C, V*), representing the vaccination strategy. Since 0 ≤ *V* ≤ 1, it is imposed the constraint *v*(*I, C, V*) = 0 ∀ (*I, C, V*) such that *C* + *V* = 1. The simplest case assumes a vaccination strategy with a constant vaccination rate for individuals, *υ* = *ϕ*, where *ϕ* is the vaccination coefficient. A more realistic representation considers that the vaccination rate is proportional to the susceptible population, *υ* = *ϕS*.

The population composed by the sum *C* + *V* constitutes the individuals that cannot become infected. Hence, neglecting the possibility of reinfection, the higher is the number of infected-vaccinated, the less is the number of susceptible individuals that can be infected. Furthermore, the absence of vaccination (*V* = 0) reduces the map to a two-population dynamics *I*-*C*.

The total number of deaths, *D*, can be estimated based on the infected population. Therefore, the cumulative number of deaths is expressed by

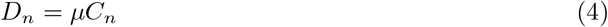

where *µ* is the death rate, usually around 2%. It should be pointed out that the current number of deaths can be determined by the difference *D*_*n*_ − *D*_*n−*1_.

The transmission rate *β* is the critical parameter to characterize the COVID-19 dynamics, being related to social isolation and virus infectious capacity. Therefore, the increase of this parameter can represent a more infectious variant for the same level of social isolation or the reduction of the social isolation for the same virus variant infectious capacity. If virus variants are neglected, the transmission rate is directly related to social isolation. In any case, the transmission rate is clearly time dependent, which motivates the definition of *β* = *β*(*n*). This time dependence can be established from an adjustment with real data, defining a proper fit. An interesting approach to match real data is the use step functions, defined as follows for an arbitraly *m* steps, given by Eq. 5 and graphically shown by Fig. 1.

**Figure 1:**
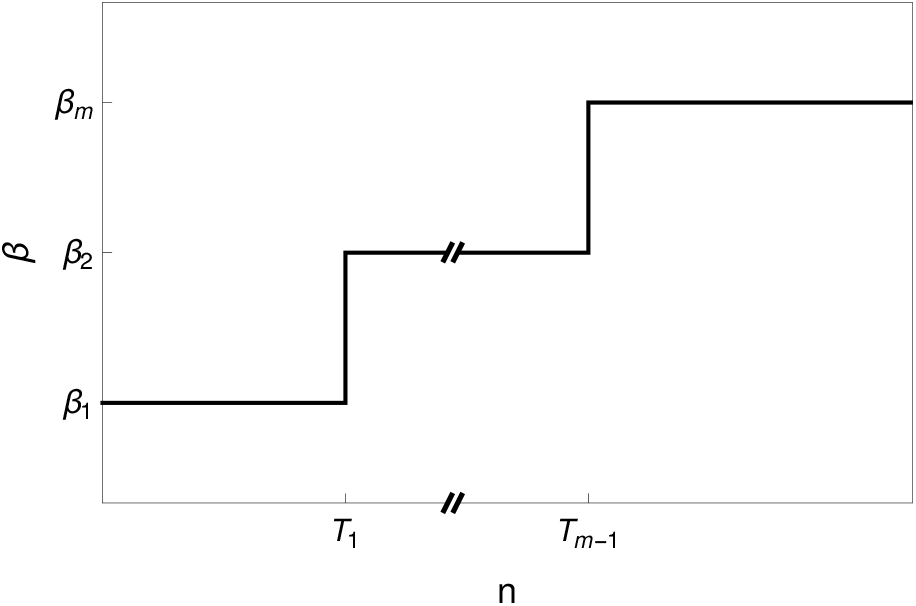
Transmission rate *β* = *β*(*n*) represented by the general step function with *m* steps

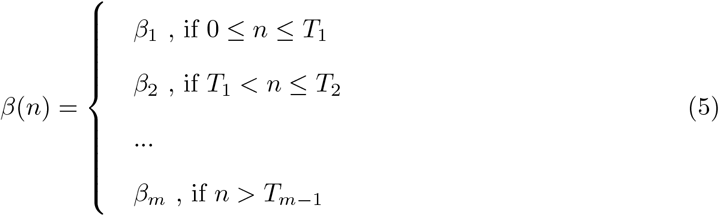

The other parameters, *σ* and *γ*, usually assumes typical values for COVID-19 dynamics [15, 17, 33]: *σ* = 1*/*3 and *γ* = 1*/*5. These parameters are employed in all simulations except when mentioned otherwise. The vaccination coefficient *ϕ* can also vary throughout time which means that some vaccination campaigns can be modeled by step functions. This approach is interesting to describe situations related to lack of vaccines, useful case for the COVID-19 pandemic.

### 2.1. Infectious Rate

The stability of the map can be evaluated from the definition of the infectious rate *r*, defined as follows based on the ratio between two subsequent iterations,

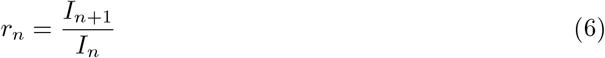

The infectious increases in a specific time if *r*_*n*_ *>* 1, and decreases otherwise, if 0 *< r*_*n*_ *<* 1. The case *r*_*n*_ = 1 is the transition between both conditions. By taking the active infected given by Eq. (2a), one can obtain the ratio *r*_*n*_ as a function of *I*_*n*_ and (*C*_*n*_ + *V*_*n*_).

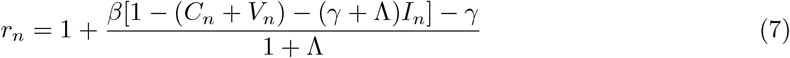

Consider the system state space *I*-(*C* + *V*) showing the curves standing for Eq. (7) with *r* = 1. This map is representative of the state space being presented in Fig. 2a for various values of *β* and constant values of *σ* and *γ*. Since the curves are related to *r* = 1, the region below each curve is associated with values of *r >* 1 while above the curve is related to *r <* 1. Therefore, the region below the curve is associated with a growth of active cases. The peak of *I* occurs when *r* = 1 is reached. Fig. 2a allows one to obtain the number of total infected plus vaccinated required to prevent the increase in the number of active cases regardless the number of currently infected and considering fixed parametric combination. In other words, when the sum *C* + *V* reaches a critical value, the infected population *I* necessarily decreases. Thus, the herd immunization point, *P*_*h*_, is defined when this critical situation is achieved and *I* = 0 (see Fig. 2b). In other words, for any given parametric combination, if *C* + *V* ≥ *P*_*h*_, then *r <* 1 ∀ *I*.

**Figure 2:**
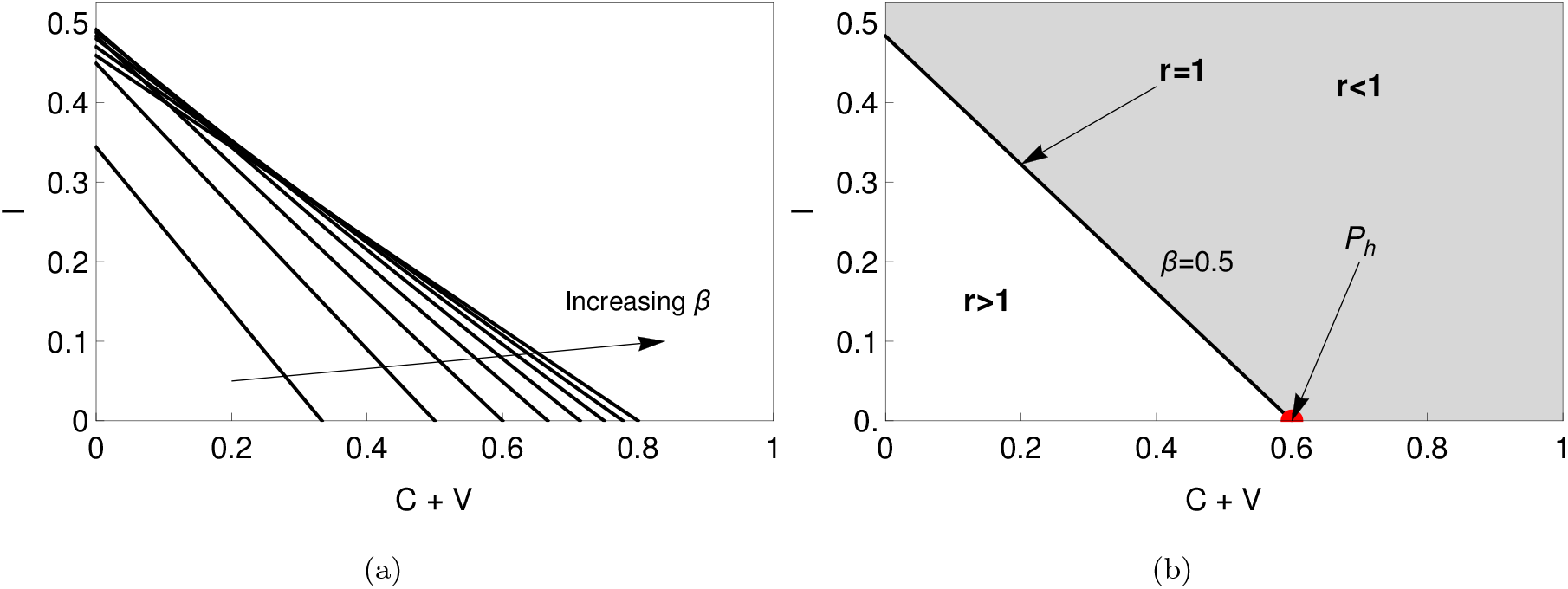
State space *I*-(*C* + *V*) with infectious rate equals to 1 (Eq. (7) with *r* = 1) for constant values of (*σ, γ*) and *β* ranging from 0.2 to 1 with step of 0.1 (a), and the herd immunization point *P*_*h*_ indication for *β* = 0.5 (b).

The value of the herd immunization point *P*_*h*_ can be analytically defined as a function of the parameters (*β, σ, γ*). By considering *r*_*n*_ = 1 and *I*_*n*_ = 0 in Eq. (7) and after some algebraic manipulation, one obtains the following expression

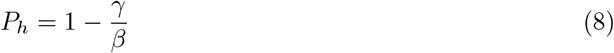

Note that this is a function of the transmission rate *β* and the infectious period *γ*^*−*1^, being not dependent on the latent period *σ*^*−*1^. Moreover, it is noticeable that an increase of the transmission rate *β* results in a higher value of *P*_*h*_. In other words, the higher is the transmission rate, the bigger is the infected-vaccinated population needed to achieve the herd immunization point. In the limit *β* → ∞, it yields *P*_*h*_ → 1, as expected. This means that, in order to have *r*_*n*_ *<* 1, it is necessary that 100% of the population becomes either infected or vaccinated. Finally, making *P*_*h*_ = 0, one obtains *β* = *γ*, which means that for any *β < γ* the number of active cases necessarily decreases regardless the number of *I* or (*C* + *V*). Therefore, the higher is the average infectious period, given by *γ*^*−*1^, the lower is the transmission rate coefficient required.

In order to present an interpretation of the COVID-19 map and the herd immunization point, consider the subspace (*I*_*n*+1_-*I*_*n*_), which is a function of (*β, σ, γ, C*), neglecting the vaccination effect. By assuming constant values of *σ* and *γ*, the map can be observed as a function of (*β, C*). Fig. 3 presents the influence of these parameters on the map curve showing a parabola that reduces its maximum value with the increase of either *β* or *C*. The dashed curve represents *I*_*n*+1_ = *I*_*n*_ that is the region where *r* = 1. Therefore, equilibrium points and their stability define the herd immunization point. Note that the fixed point 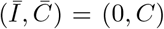 is stable if *dI*_*n*+1_*/dI*_*n*_ *<* 1. Thus, the threshold point *P*_*h*_ is calculated for *dI*_*n*+1_*/dI*_*n*_ = 1, which results in

**Figure 3:**
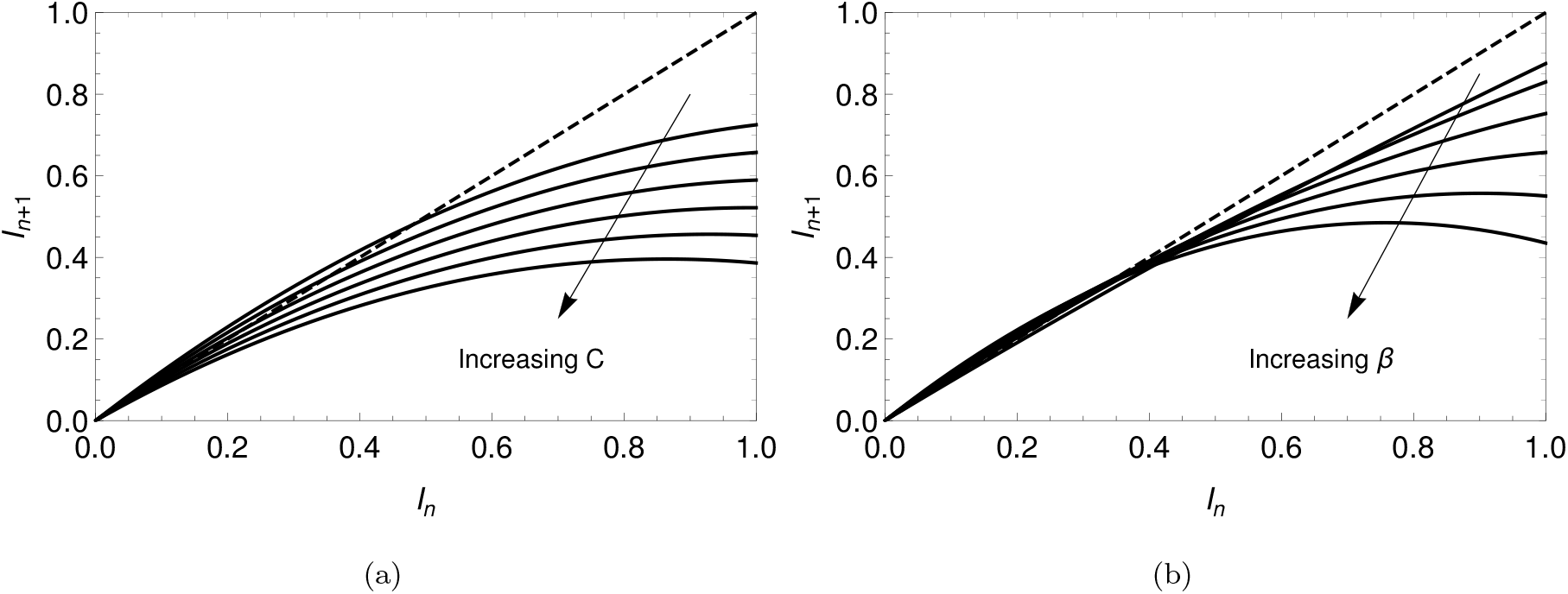
Map *I*_*n*+1_-*I*_*n*_ for *β* = 0.8 and varying *C* from 0 to 1 with a step of 0.2 (a); and with constant *C* = 0.2 and varying *β* from 0.2 to 1.2 with step of 0.2 (b). The dashed line stands for *I*_*n*+1_ = *I*_*n*_.

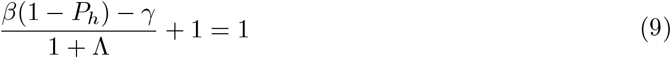

that is the same result of Eq. (8),

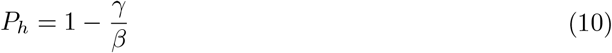

The stability can be observed from Fig. 4 that shows two different scenarios employing different parameters. The first scenario is related to an unstable case with *β* = 0.5, and the other one is a stable case with *β* = 0.05. Different initial conditions are of concern in order to illustrate the system evolution.

**Figure 4:**
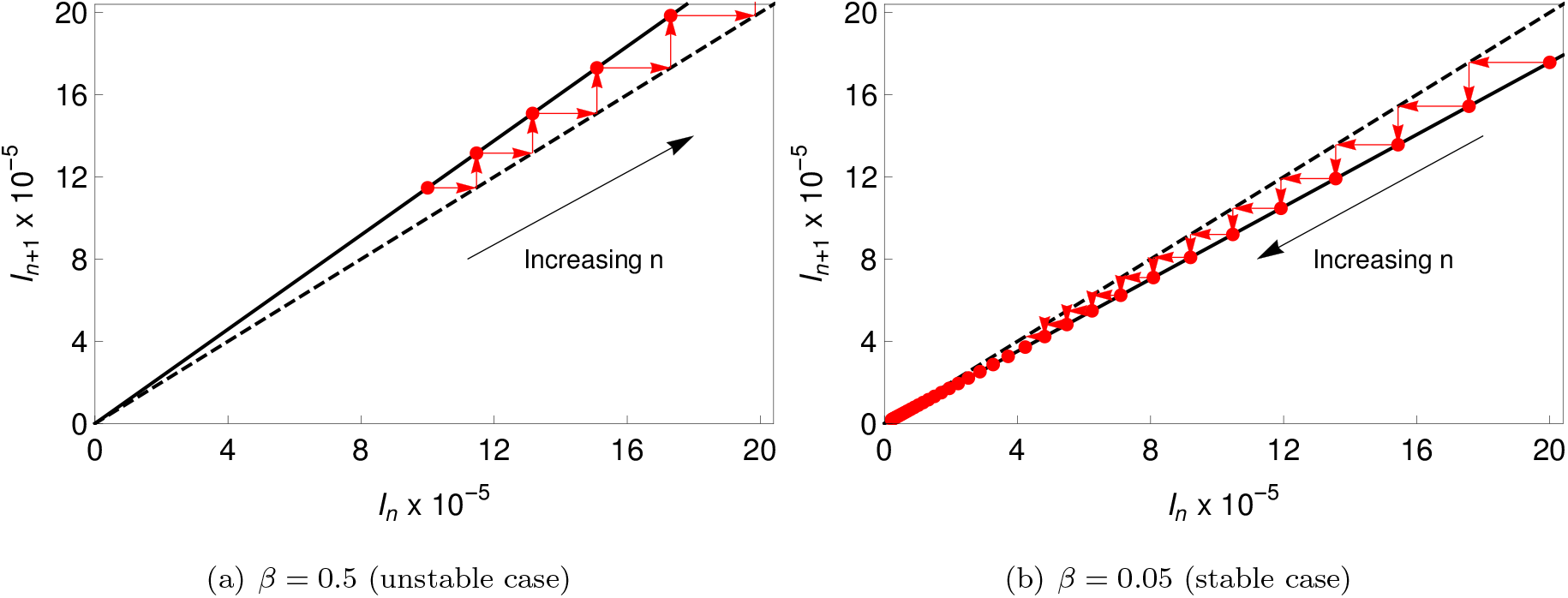
Map (*I*_*n*+1_-*I*_*n*_) for *C* = 0 showing an unstable (a) and stable (b) cases. The dashed line stands for *I*_*n*+1_ = *I*_*n*_.

In the sequence, the COVID-19 map is applied to describe the epidemic dynamics using real data from Germany, Italy and Brazil as references.

## 3. Model Verification

The novel COVID-19 map is now employed to perform a dynamical analysis of the pandemic. Real data from the novel COVID-19 epidemics in Germany, Italy and Brazil are employed as reference considering information from Worldometer (https://www.worldometers.info/). The comparison is established by considering nondimensional values, dividing each population by the total population of the country. In this regard, the following values are adopted to each country: Germany *N*_ger_ = 83.03 × 10^6^; Italy *N*_ita_ = 60.36 × 10^6^; Brazil *N*_bra_ = 211 × 10^6^. The time period employed ranges from 6th March of 2020 to 21st January of 2021. Due to natural seasonality in real data, a 7 day average is employed, giving rise to a new data set. Both infected *I* and cumulative *C* populations are of concern (Fig. 5) showing a two-wave pattern. For Germany and Italy cases, the first wave peak occurred after April 2020, while the Brazilian case shows a longer first wave, which is actually a plateaus pattern. Therefore, the Brazilian second wave started from a high level value. On the other hand, Germany and Italy present the second wave peak around December 2020 and the number of active cases are droping at the end period. Based on these observations, COVID-19 is characterized by different patterns. A bell shape behavior is the essential point to be observed, but it is clear the possibility of either multi-wave or plateaus patterns.

**Figure 5:**
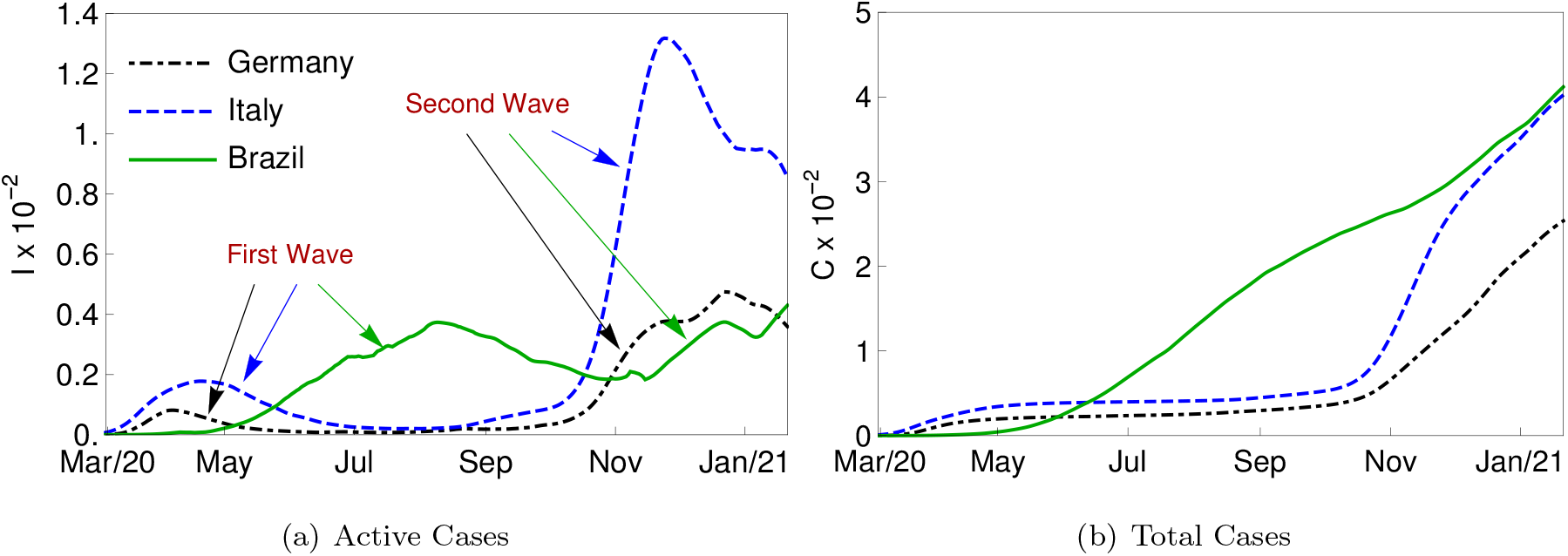
Active (left) and Total (right) cases of the novel coronavirus for Germany (black), Italy (blue) and Brazil (green).

COVID-19 map is able to represent the bell shape behavior by considering proper parameters. Based on this, consider a limit period of the real data given by the first 120 days. Fig. 6 presents results of the map simulations compared with real data using the least square method (LSM) for the fitting process. It is noticeable a good agreement between numerical and real data. Transmission rate is described by step functions adjusted for each country: *β*_ger_ for Germany, *β*_ita_ for Italy and *β*_bra_ for Brazil. These functions are displayed in Eq. (11), where *n* = 0 yields 6th March 2020.

**Figure 6:**
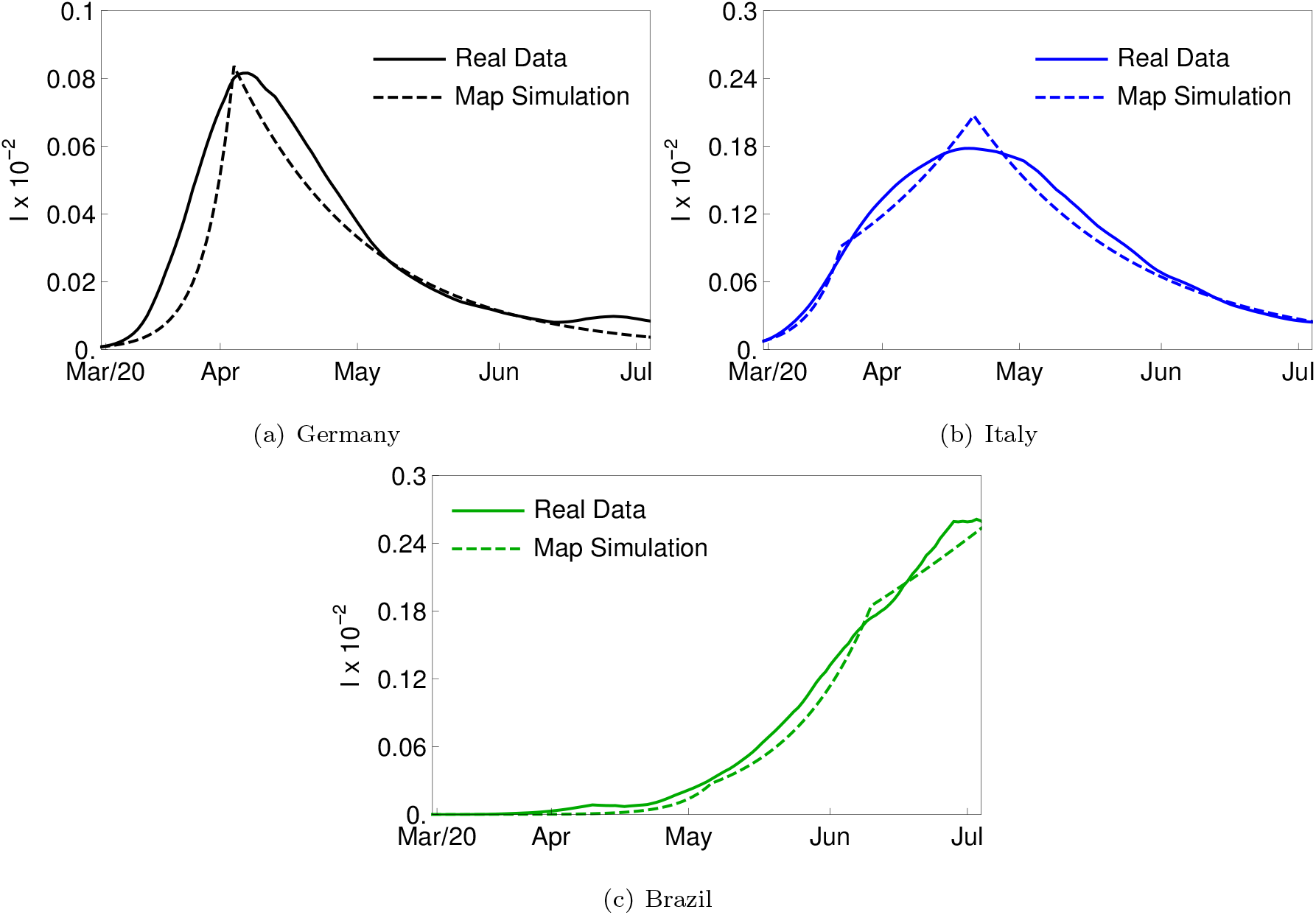
Infected active cases *I* for Germany (a), Italy (b) and Brazil (c) for the first 120 days of the period range employed. The continuous line stands for real data and dashed line for the map simulation, which employed *σ* = 1*/*3, *γ* = 1*/*5 and *β* furnished by Eq. 11.

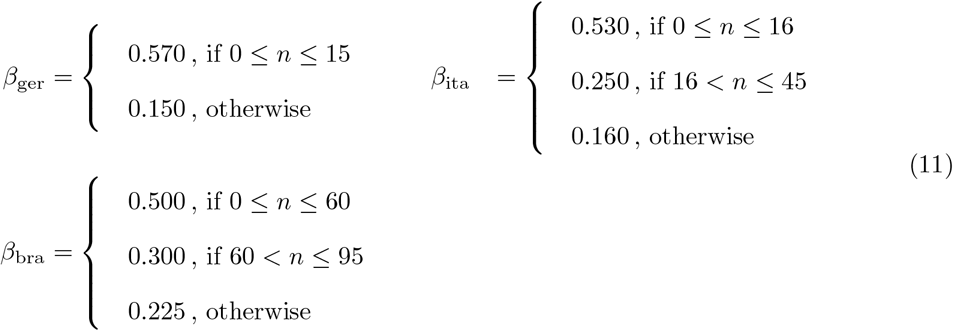

In addition to real data, model verification establishes a comparison of the COVID-19 map with the SEIRV model that is integrated employing the fourth-order Runge-Kutta method with a 10^*−*2^ time step. The vaccination effect is neglected in this stage, adopting *V* = *υ* = 0. Analytic considerations and an explicit discrete-continuous comparison are of concern.

Due to the bell shape characteristics of active cases, three different aspects characterize an outbreak: the peak of the active cases curve, *I*_max_; the time instant where the peak occurs, *t*_max_; and the area below the curve, which is proportional to the total infected after the outbreak *C*(*t* → ∞). This latter characteristic can be confirmed by analyzing the SEIR model, substituting Eq. (1e) into Eq. (1c) and integrating from the beginning of the outbreak to its end, which gives

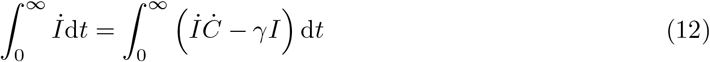

and therefore,

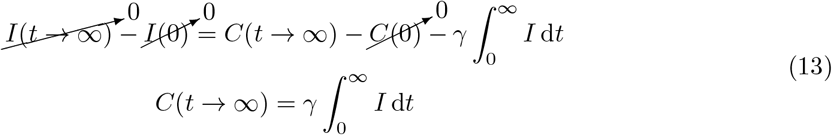

These three aspects can be used to build the model signature in 3D charts (*I*_max_ - *t*_max_ - *C*(*t* → ∞)). The variation of the three parameters (*β, σ, γ*) generates a solid object in the 3D chart that can be understood as the model signature.

In order to facilitate the signature view, the 3D chart is split into two 2D maps. Additionally, *σ* and *γ* are assumed to be constant and *β* is free to vary. On this basis, a curve belonging to the model signature is obtained characterizing a scenario. Four different combinations for (*σ, γ*) are picked to compare the novel map and the SEIR model, as presented in Figs. 7 and 8 considering *C*_0_ = *I*_0_ = 10^*−*5^. In general, it is noticeable that the signature predicted by both models are in close agreement considering the total cases against *t*_max_. This convergence takes place regardless the parametric condition. On the other hand, Fig. 7 shows that the signature of both models diverge for bigger values of *β*, being the map less sensitive to *β* variation. Besides, the same effect is obtained with the increase of *γ*^*−*1^. Nevertheless, there is a region where both models diverge from each other. This divergence does not imply that both models cannot be used to model the evolution of an outbreak, representing just a quantitative divergence. It should be pointed out that both models present the same trend for parametric variation. Moreover, within the convergence region, the same *β* value does not necessarily generate the same combination (*I*_max_, *t*_max_, *C*(*t* → ∞)) for both models. Therefore, there is a good qualitative agreement between the novel map and the classical SEIR model.

**Figure 7:**
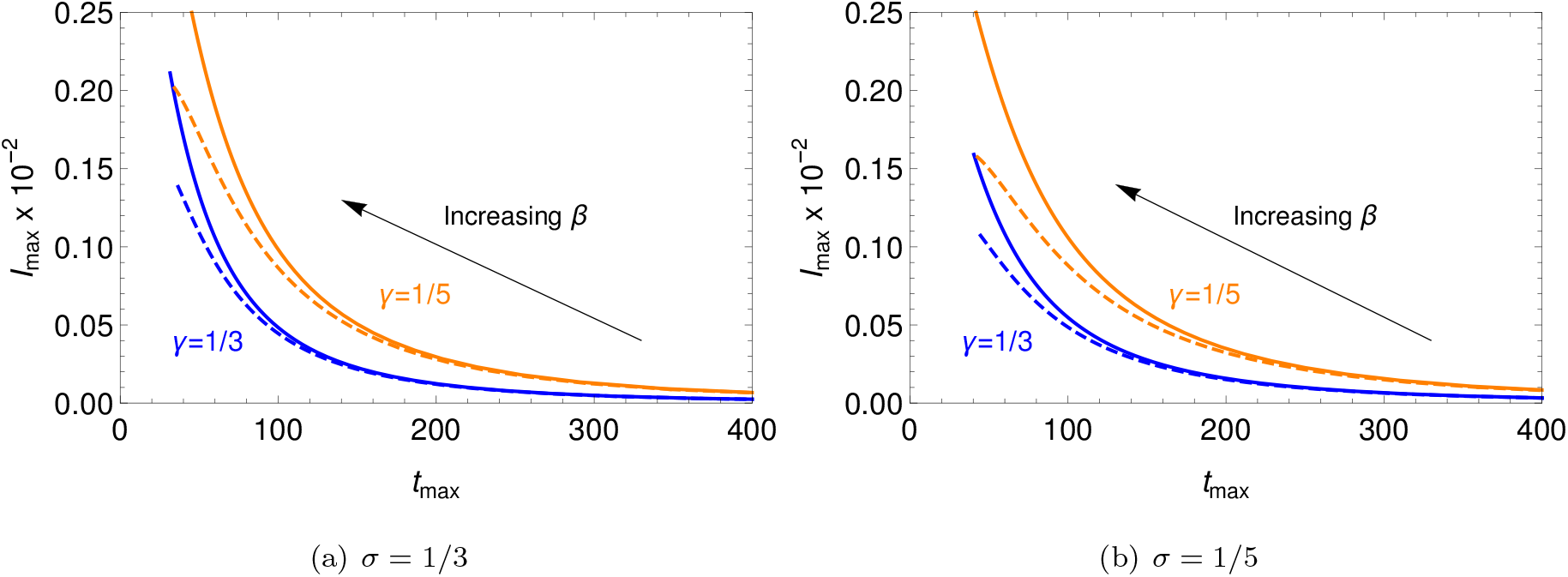
Comparison among SEIR (continuous line) and discrete map (dashed line) models for *I*max against *t*max. *β* ranged from *γ* to 1.5.

**Figure 8:**
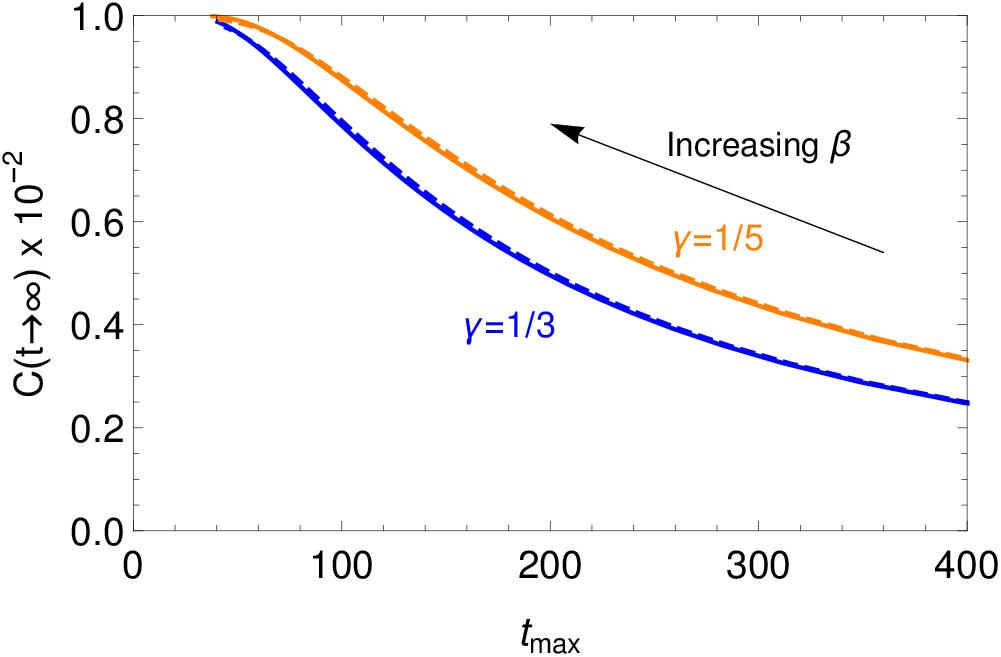
Comparison among SEIR (continuous line) and discrete map (dashed line) models for *C*(*t → ∞*) against *t*max. The total infected are independent of *σ* parameter in both models. *β* ranged from *γ* to 1.5.

The influence of the parameters on the map dynamics is now of concern. Fig. 9 presents the evolution of the outbreak simulation through the active cases *I* and the total cases *C* for different parameters. Fig. 9a presents the influence of transmission rate *β* showing that the peak of active cases is bigger and takes place sooner with the increase of the transmission rate. Additionally, the total number of infected increases as *β* increases. The influence of the mean latent period *σ*^*−*1^ is presented in Fig. 9b showing that the higher is the mean latent period, the later the peak of active cases takes place and the smaller is the peak reached by *I*. The total cases reached the same vale regardless of the value of *σ*. The variation of the latent period can change the course throughout time of *I*, but it does not influence the total number of infected. Finally, Fig. 9c presents the influence of the mean infectious period *γ*^*−*1^ on the outbreak evolution. The longer is the infectious period, the more time is available to someone infected to spread the virus among the others. Hence, the peak of active cases is higher and the total cases *C* is higher as well.

**Figure 9:**
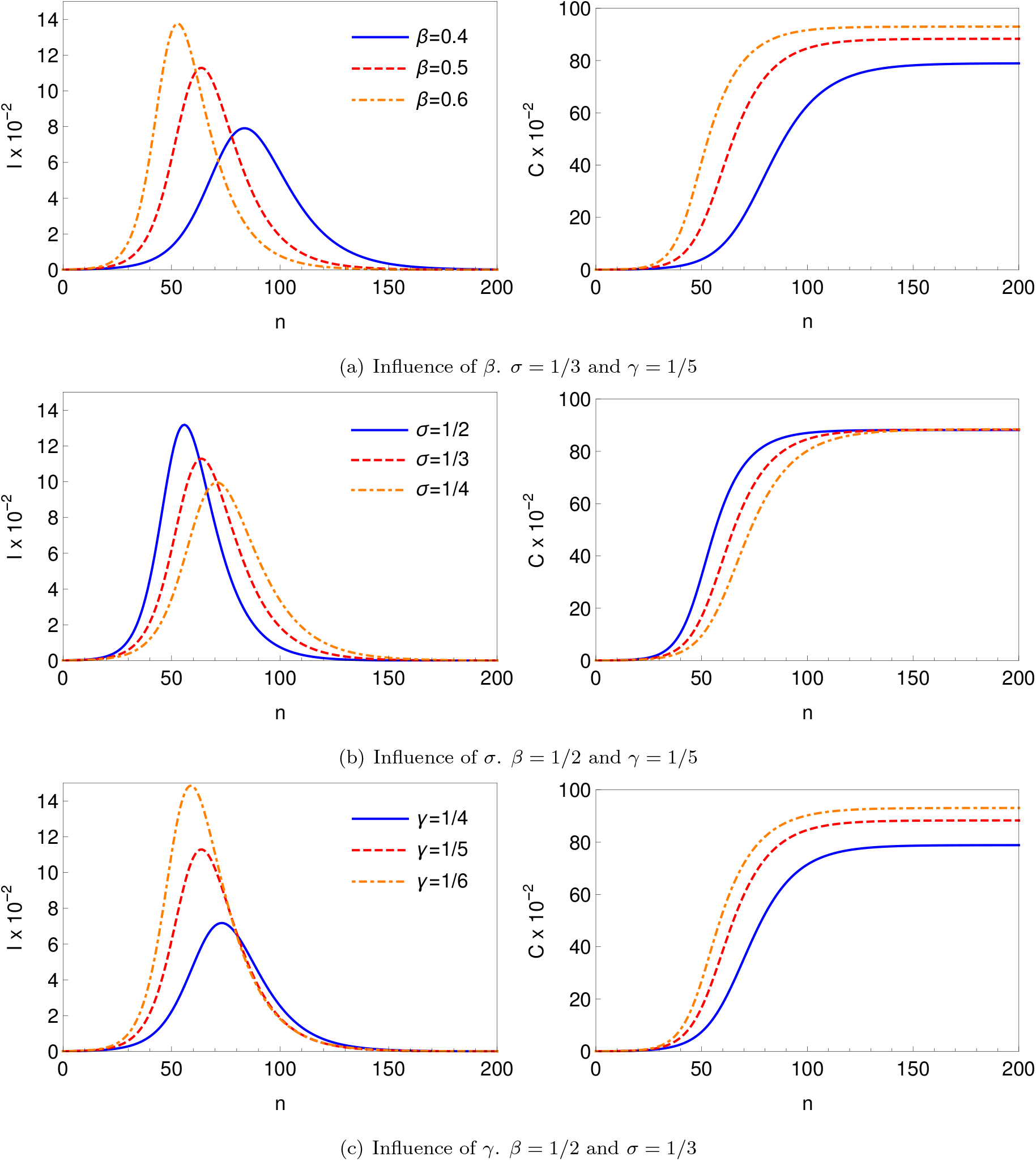
Outbreak simulation for different parametric combination. All simulations are carried out with *C*_0_ = *I*_0_ = 10^*−*4^.

### 3.1. Multi-wave Scenarios

Based on epidemic real data, it is clear that COVID-19 dynamics has a multi-wave bell shape pattern. The previous section presented results showing the capability of the map to describe a single bell shape. This section is dedicated to evaluate the multi-wave pattern. Variations of the transmission rate is the most important parameter to capture this multi-wave behavior. In this regard, consider a hypothetical epidemic scenario where *β* is described by the following step function, illustrated by Fig. 10.

**Figure 10:**
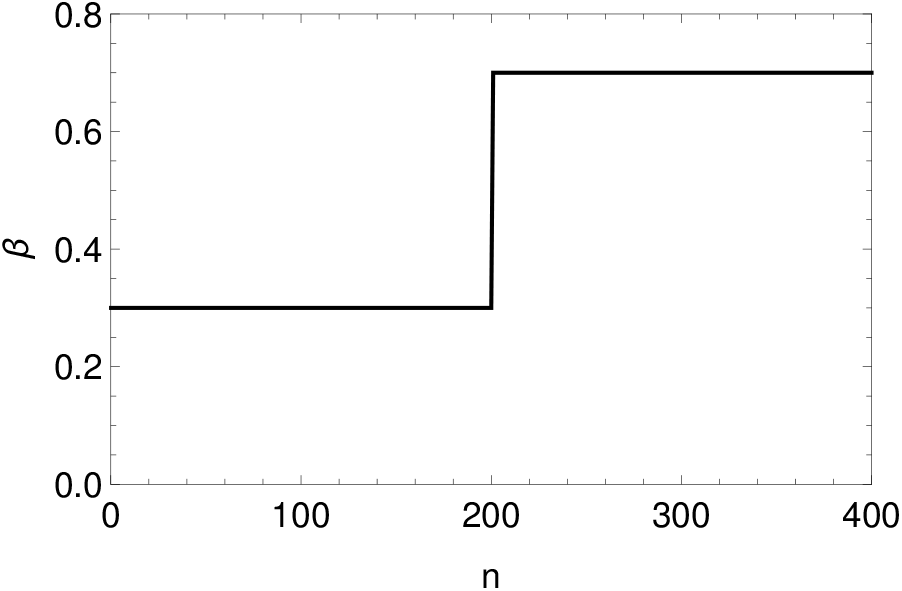
Step function of *β*

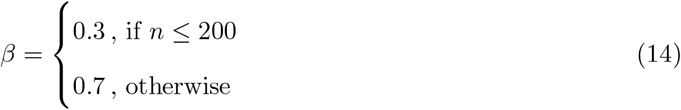

By considering COVID-19 parameters and neglecting vaccination effect (*V* = *υ* = 0), with initial conditions *C*_0_ = *I*_0_ = 10^*−*4^, the active and total cases vary with time according to Fig. 11. Results show two subsequent waves with bell shape characteristics. By observing the state space *I*-*C*, depicted by Fig. 12, it is noticeable that the number of active cases *I* begins to drop when *r* = 1 for *β* = 0.3. The abruptly increase of *β* from 0.3 to 0.7 leads to another wave and the increase of *I*. Due to the parameter change, the curve on the state space *I*-*C* for *r* = 1 changes its position and this new condition defines the onset of the second reduction of the number of active cases.

**Figure 11:**
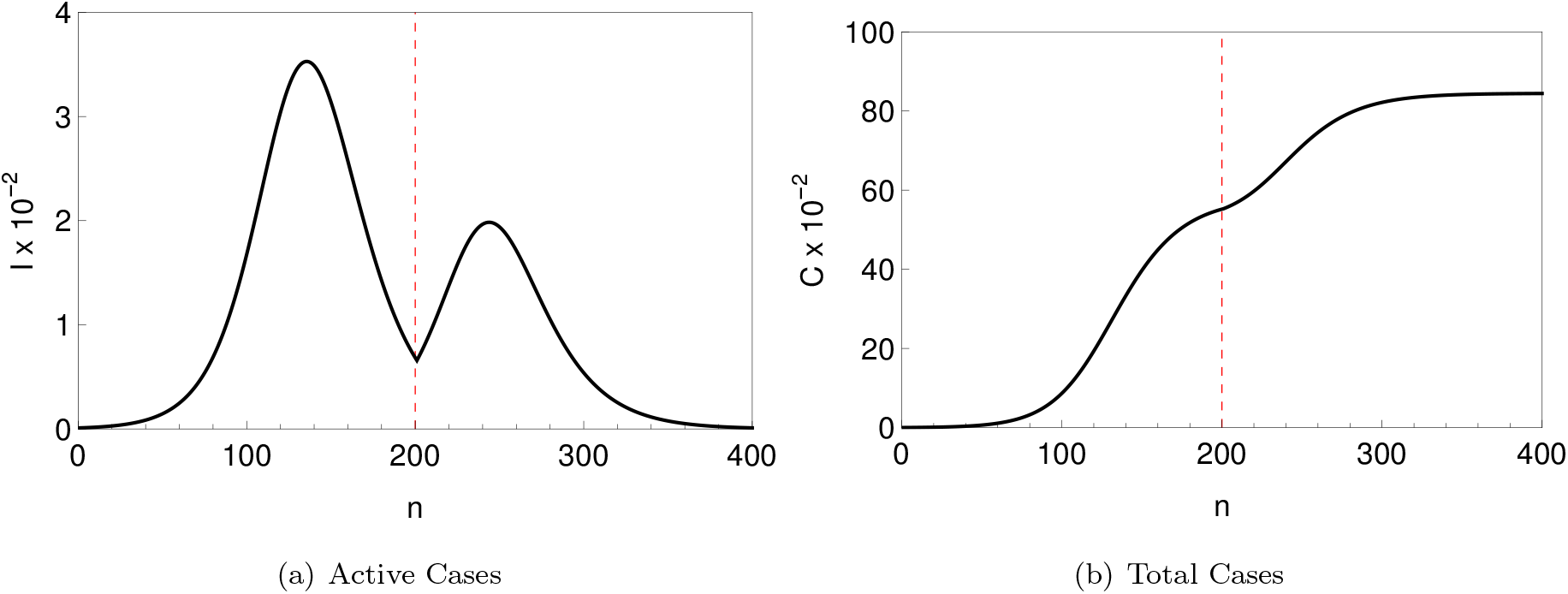
Active (left) and total (right) cases along time. The red dashed line yields *n* = 200.

**Figure 12:**
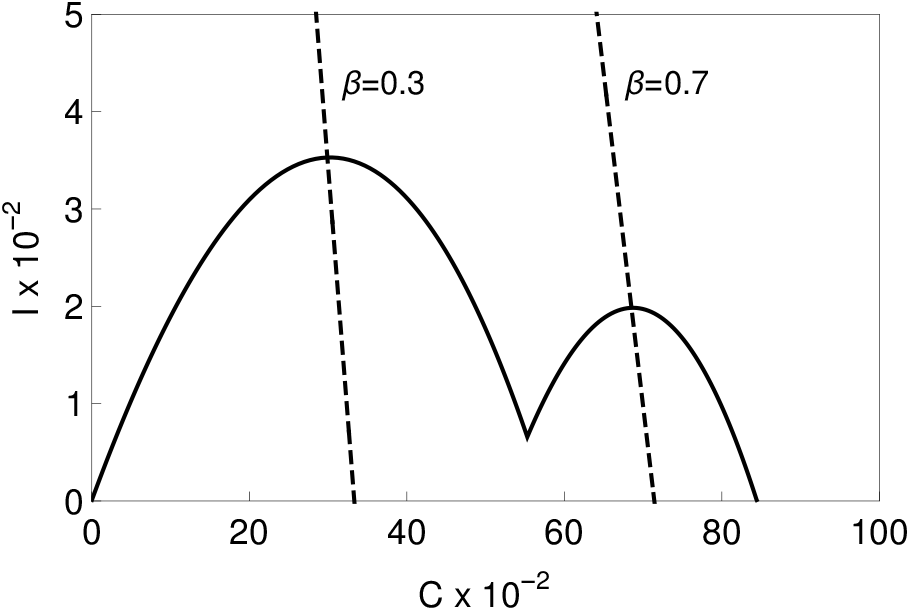
Outbreak evolution ploted in the state space. The dashed lines stand for Eq. (7) with *r* = 1 and for *β* = 0.3 and 0.7.

After the two waves, the total infected cases reached *C*(*n* → ∞) = 0.841. By using this value as the herd immunization point into Eq. (8), one obtains *β* = 0.762. Therefore, if the transmission rate is changed again, it only leads to a third wave if *β >* 0.762. Otherwise, the spread of virus is under control.

This strategy is now employed to verify the capability of the COVID-19 map to represent real data. Once again, COVID-19 epidemic in Germany, Italy and Brazil are employed as reference. The comparison among numerical simulations and real data is now carried out for the whole period range employing the least square method to perform adjustment. Transmission rate is defined by step functions presented in Eq. 15. Fig. 13 presents the comparison between numerical simulations and real data showing a good agreement for all countries. Based on that, it is possible to conclude that the COVID-19 map captures real data including the multi-wave scenario. The main difficulty is the proper determination of the transmission rate.

**Figure 13:**
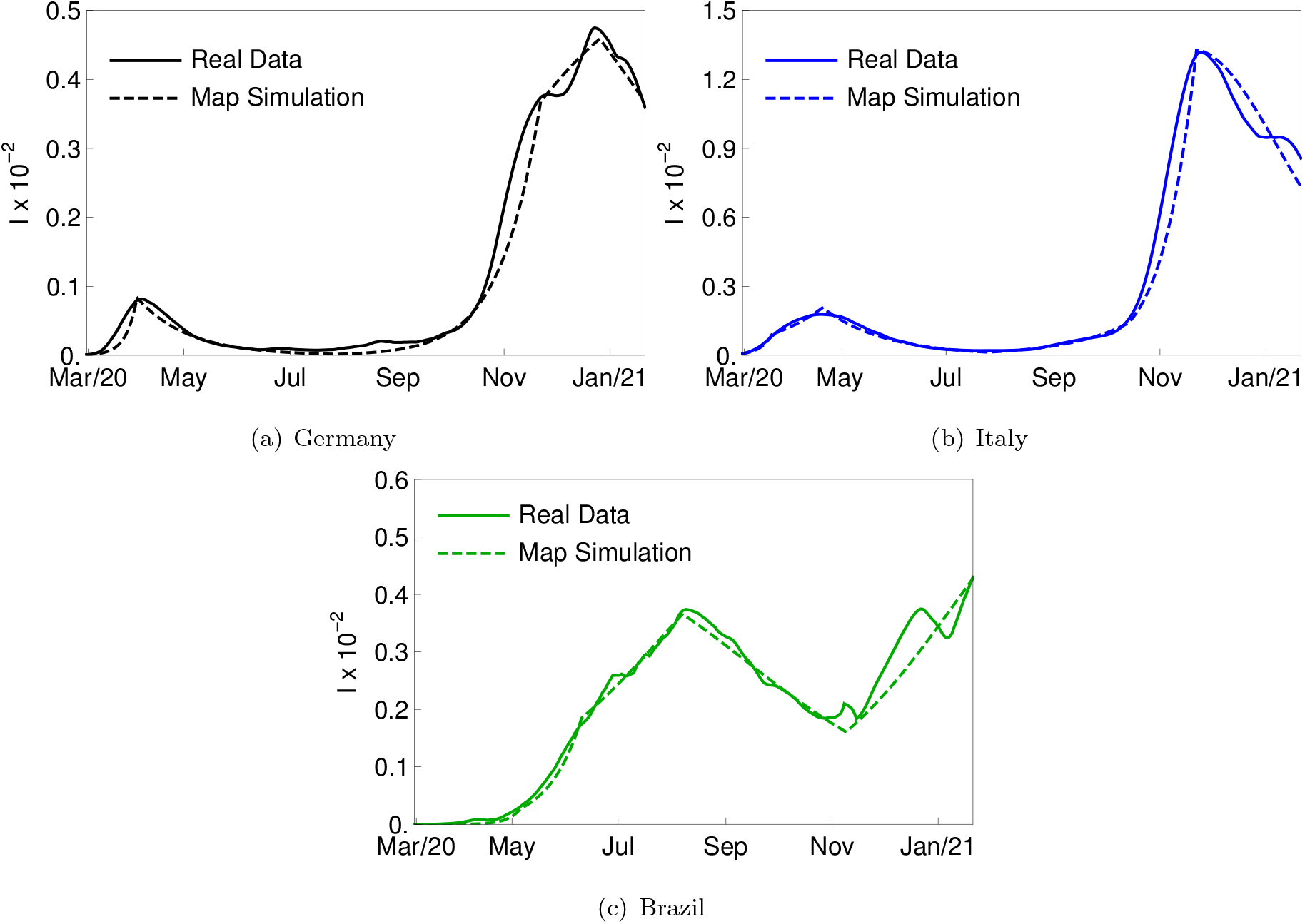
Infected active cases *I* for Germany (a), Italy (b) and Brazil (c) for the whole period range employed. The continuous line stands for real data and dashed line for the map simulation, which employed *σ* = 1*/*3, *γ* = 1*/*5 and *β* furnished by Eq. 15.

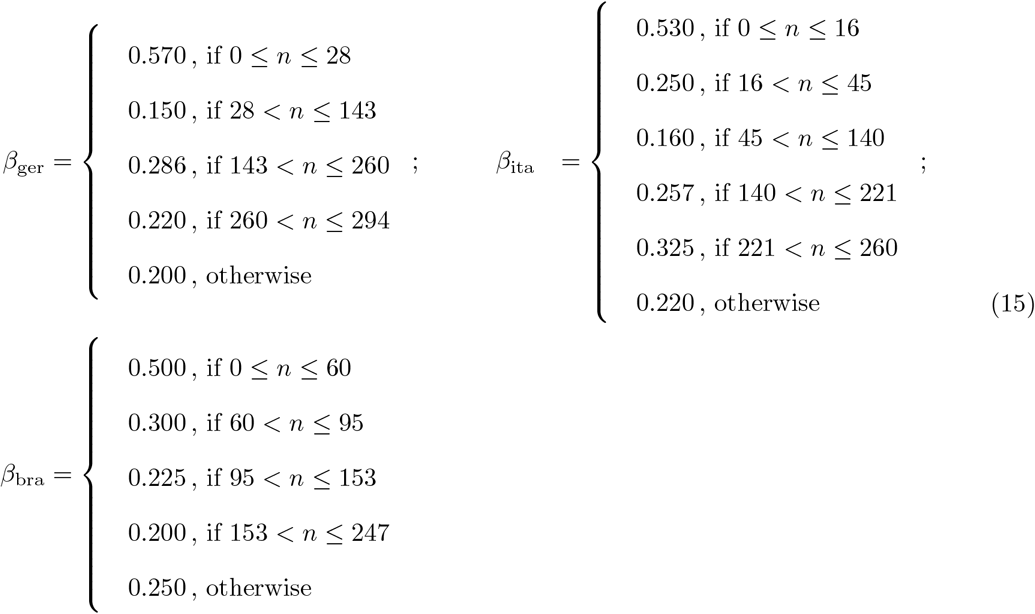

## 4. COVID-19 Dynamical Analysis

The novel COVID-19 map is now employed to perform a dynamical analysis of the pandemic. As mentioned before, the transmission rate is the critical point for a proper description of the pandemic evolution. An approach to represent the transmission rate is to define a function of time *β* = *β*(*n*) that is adjusted from real data employing Eq. (2a). Based on real data time series, the Newton’s method is employed to calculate *β*(*n*) at each time step for each country. Fig. 14 presents the estimated transmission rate, *β*(*n*), together with the infected active cases *I* showing that the real data is reproduced accordingly. It is observed that transmission rate are bigger during the first part of the outbreak. The reduction that follows is associated with social isolation policies. It should be pointed out that the growth of active cases are related to periods where *β* is bigger than ≈ 0.2, the value estimated by considering *P*_*h*_ = 0. This is more explicit during the first and second epidemic waves where it is possible to observe the increase of the cases.

**Figure 14:**
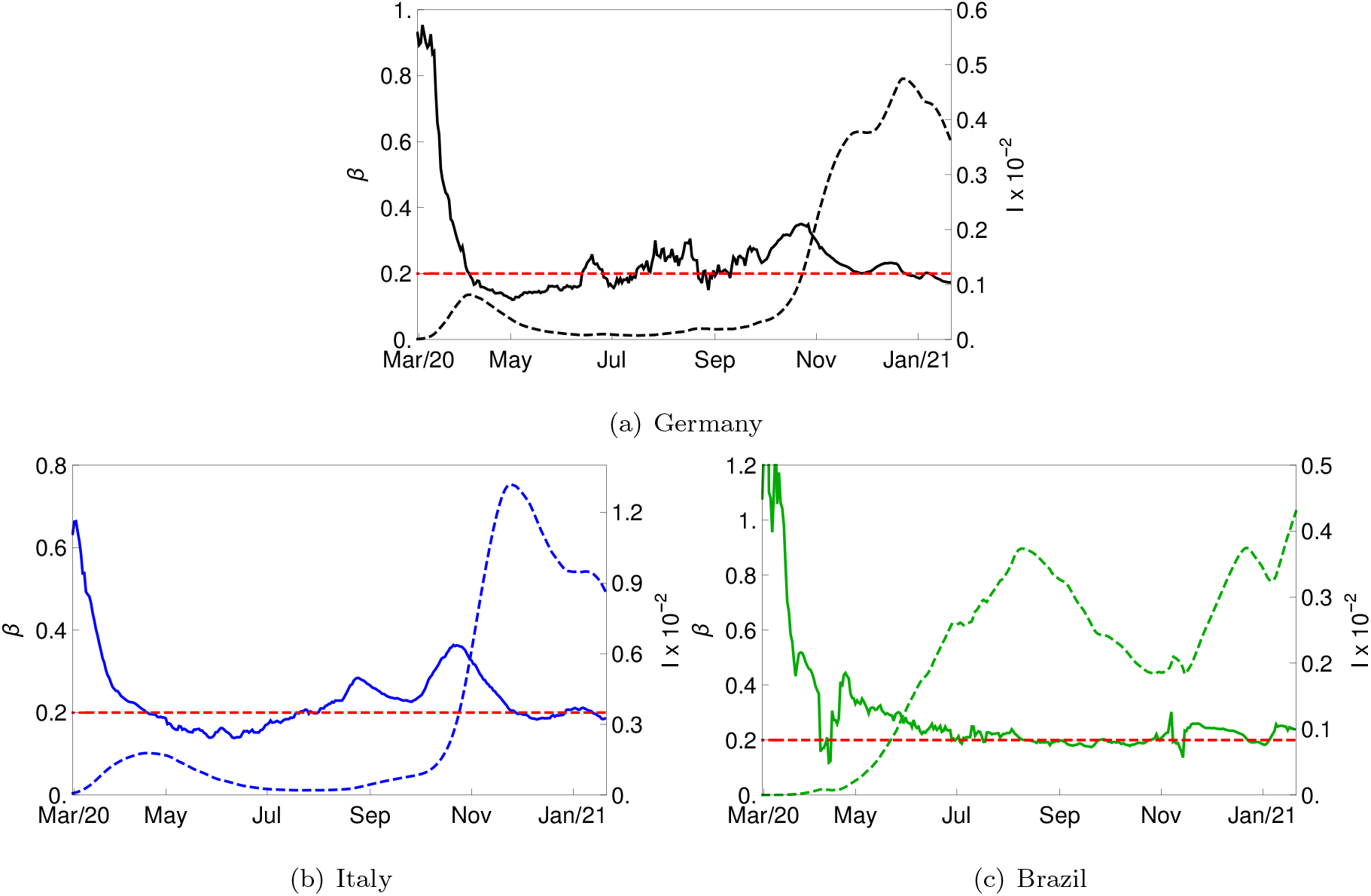
Transmission rate *β* (continuous line) ploted with active cases *I* (dashed line) for each country. The red dashed line stands for *β* = *γ* = 0.2 for the herd immunization point *P*_*h*_ = 0.

An extrapolation for future values are now of concern in order to evaluate the outbreak evolution on the state space *I*-*C*. The critical transmission rates is obtained for *r* = 1 (employing Eq. (8)) at the last day of the period, yielding *β*_*cr*_ = 0.206 for Gemany, 0.210 for Italy and 0.209 for Brazil. Fig. 15 presents the map simulation *I*-*C* for the three countries, highlighting the curve for the critical infectious rate *r* = 1 with *β* = *β*_*cr*_ and showing different outbreak simulations starting from the last day of the period, for each country, associated with distinct scenarios defined by constant values of the transmission rate *β*. The curve yielding *r* = 1 for *β* = *β*_*cr*_ is shown as reference. Note that transmission rate values below *β*_*cr*_ leads to the reduction of the number of active cases. Otherwise, if the transmission rate is bigger than *β*_*cr*_, the cases tend to increase. On this basis, transmission rate is the essential parameter to control the pandemic evolution, being possible to define the critical situation.

**Figure 15:**
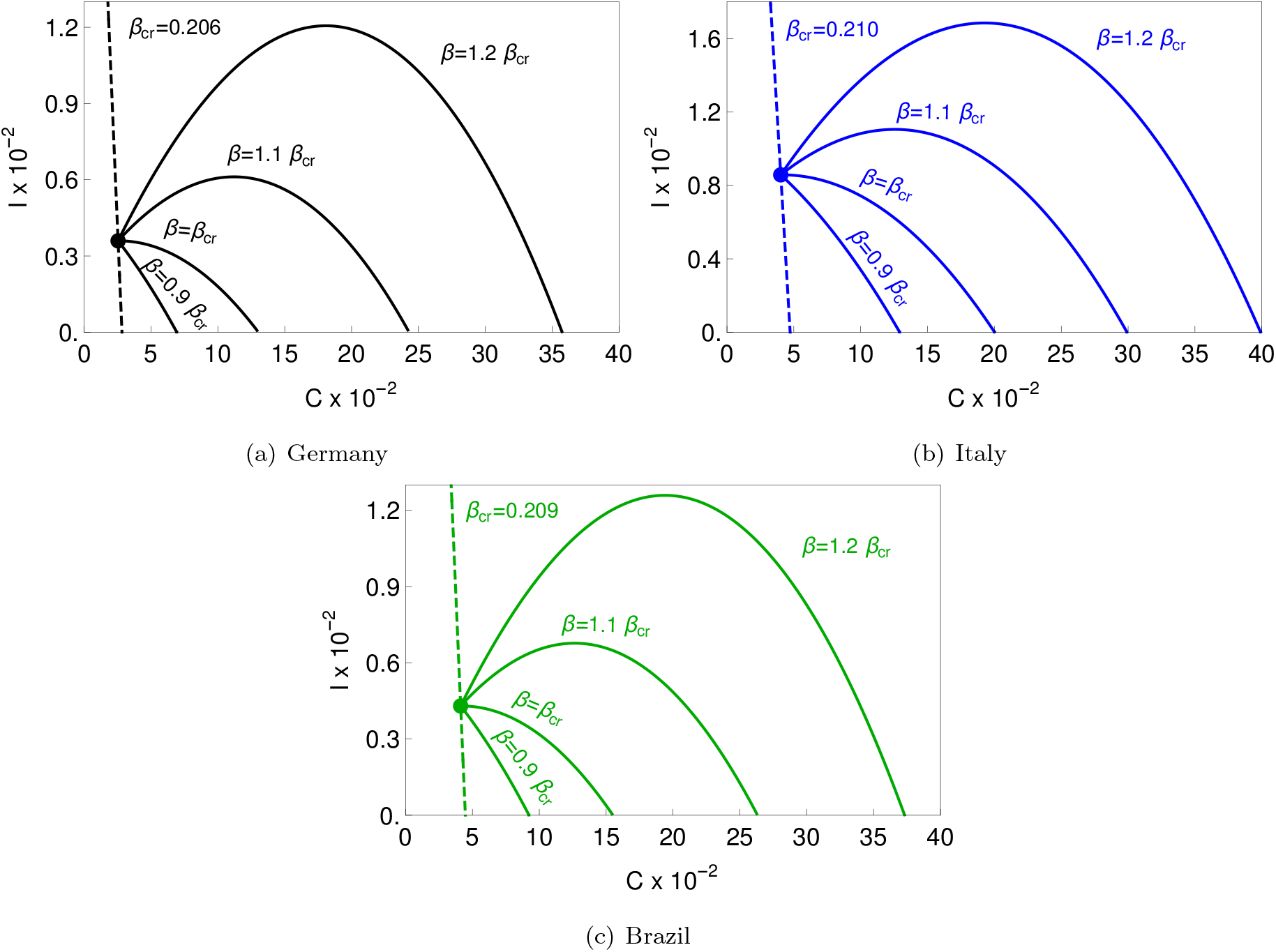
COVID-19 dynamics on the map *I*-*C* for each country (continuous line). The dashed line stands for simulations from the last day period carried out for various fractions of *β*_*cr*_. The dashed line stands for the infectius rate *r* = 1 with *β* = *β*_*cr*_.

The analysis of deaths from COVID-19 infection is now of concern. The death ratio *µ* stands for the ratio between total cases and total deaths. Since the number of cases is directly associated with the transmission rate, it is expected that this ratio is a function of time, *µ* = *µ*(*n*). Fig. 16 presents *µ* adjusted from real data considering Germany, Italy and Brazil. Note that this ratio varies through time showing that at the beginning it assumes bigger values. As time goes by, values evolve to lower values, probably due to the development of healing strategies. The average of *µ* for the last 90 days of the period range is 1.86% for Germany, 3.95% for Italy and 2.68% for Brazil. These values are employed in the sequence in order to evaluate the vaccination effect.

**Figure 16:**
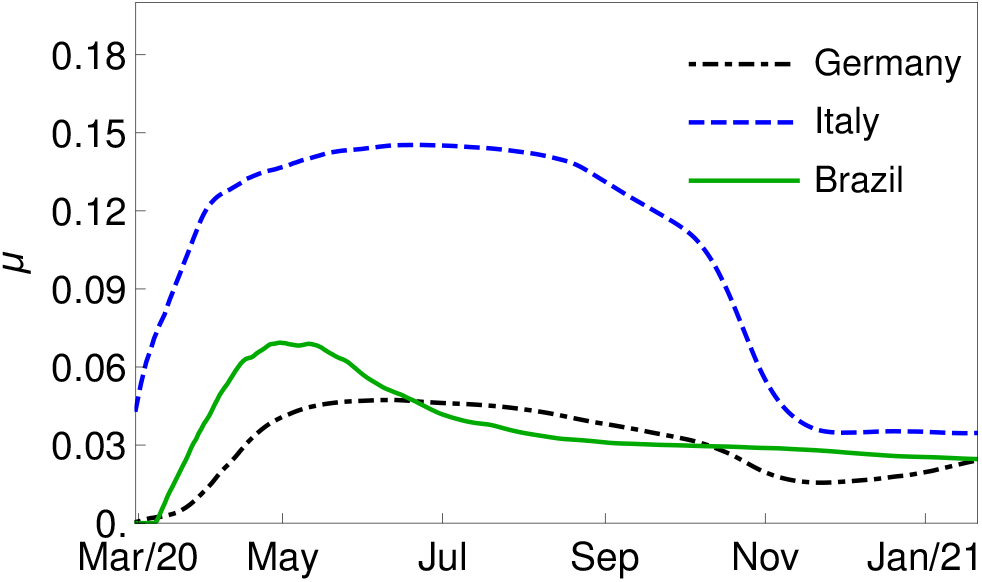
Death ratio *µ* along time for each country.

The relationship between transmission rate and herd immunization can be established by considering the number of total infected required to maintain the infectious rate less than one (*r <* 1). Eq. (7) can be used in order to build the herd immunization threshold point *P*_*h*_ as a function of *β* (Fig. 17). The average of transmission rate values during the first seven days of the period range is 0.91 for Germany, 0.62 for Italy and 1.26 for Brazil. Based on these values, the herd immunization point *P*_*h*_ is achieved only with total infected cases *C* above 0.78, 0.68 and 0.84, respectively, as depicted in Fig. 17. This scenario suggests the only way to avoid a high number of total infected without the need to maintain social isolation is employing population vaccination. Therefore the sum *C* + *V* can be high enough in order to allow a high *β* coefficient. The effect of vaccination is explored in the following section.

**Figure 17:**
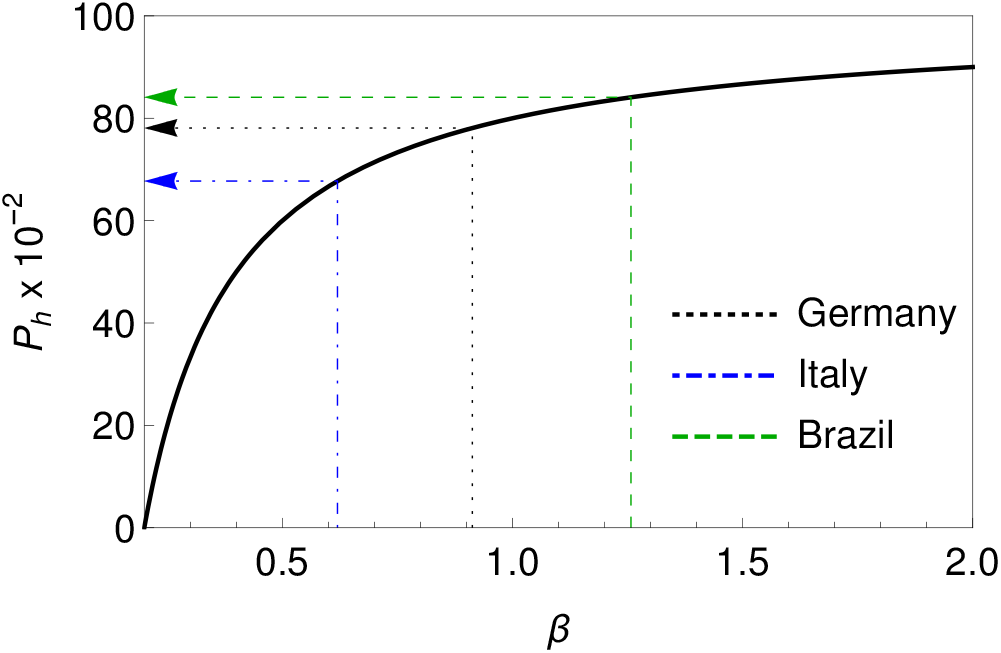
Herd immunization *P*_*h*_ as a function of transmission rate *β* (Eq. (8)) with *γ* = 1*/*5 (COVID-19 parameter). The dotted (Germany), dot-dashed (Italy) and dashed (Brazil) lines indicate *P*_*h*_ required for *β* = 0.78, 0.68 and 0.84, respectively for Germany, Italy and Brazil.

## 5. Effect of Vaccination

This section investigates the effect of vaccination on the COVID-19 dynamics. It is adopted a situation where the vaccination starts on the last day of the real data period: 21st January 2021. The transmission rate employed is the mean value from *β*(*n*) of the last 90 days of the period range (see Fig. 14) which yields: *β*_ger_ = 0.227, *β*_ita_ = 0.230 and *β*_bra_ = 0.224. Moreover, three vaccination scenarios are analyzed with the following coefficients: *ϕ* = 0, 10^*−*3^ and 10^*−*2^, where the case *ϕ* = 0 stands for no vaccination. Besides that, two vaccination campaign models are treated: *υ* = *ϕ* and *υ* = *ϕS*. On this basis, the vaccinated population *V* is described by one of the following equations:

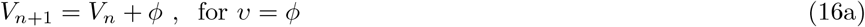

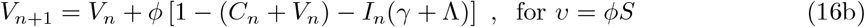

It should be pointed out that either Eq. 16a and 16b can be combined with Eq. 2a and 2b, which reduces the dynamical system to two independent equations, associated with variables (*I, C* + *V*). Based on that, Fig. 18 presents a comparison between these two vaccination models for two vaccination coefficients and two transmission rates. An essential characteristic of the dynamics is represented by the peak time instant, *t*_max_, and the difference *C*(*t* → ∞)−*C*_0_. Note that a low value of transmission rate *β* (Fig.18a) causes that both models yield approximately the same outbreak evolution for the two carried out values of *ϕ*. The increase of *β* promotes a relevant difference between these vaccination models, depending on initial conditions *C*_0_ + *V*_0_. Concerning the herd immunization point, one should notice that *t*_max_ → 0 when *C*_0_ + *V*_0_ → *P*_*h*_ for all *ϕ* and *β* and for both models.

**Figure 18:**
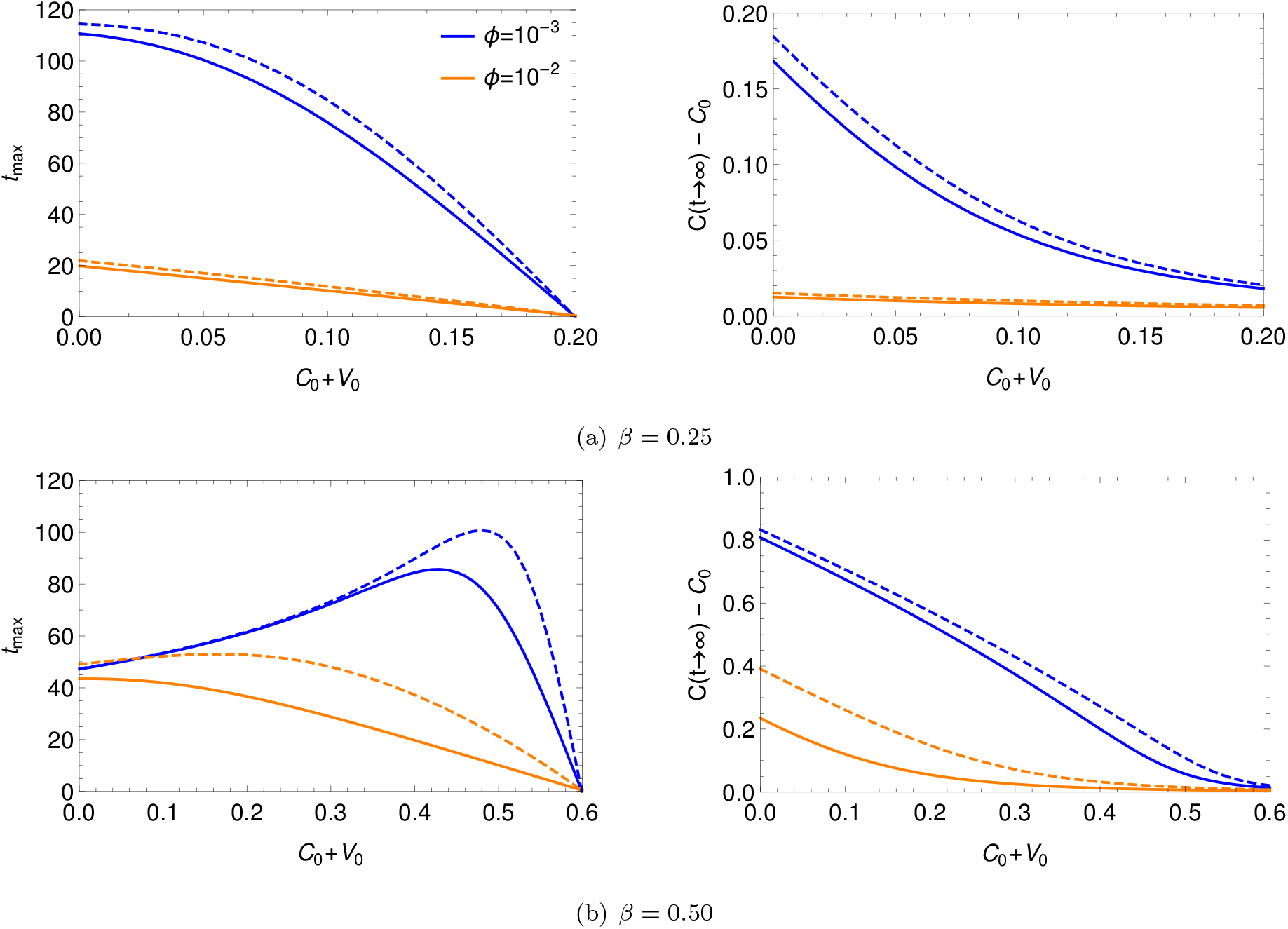
Comparison among different vaccination strategies: *υ* = *ϕ* (solid line) and *υ* = *ϕS* (dashed line). Herein, it was employed *I*_0_ = 10^*−*3^ and COVID-19 parameters.

Based on this analysis, it should be pointed out that the vaccination models generate approximately the same epidemic response. Therefore, for the sake of simplicity, the simplest model is employed to represent the vaccination effect: *υ* = *ϕ*.

Fig. 19 presents the effect of vaccination on the evolution of the epidemics for each country, showing that vaccination has a huge impact on the COVID-19 dynamics. The higher the vaccination rate is, the lower is the peak reached by active cases and the lower is the total infected population after the epidemic period. Moreover, it should be noticed that the time required to achieve, for instance, *I* = 10^*−*4^ - one infected individual for each ten thousand inhabitants, takes place sooner for higher vaccination rates. As expected, the absence of vaccination results in the worst scenario. These conclusions can be drawn for all the three countries. Fig. 20 evaluates the vaccination effect by the map *I*-(*C* + *V*). The curve standing for infectious rate *r* = 1 is also plotted, yielding the peak reached in active cases. An important conclusion is that vaccination is the only possibility to make *P*_*h*_ increase without increasing *C*.

**Figure 19:**
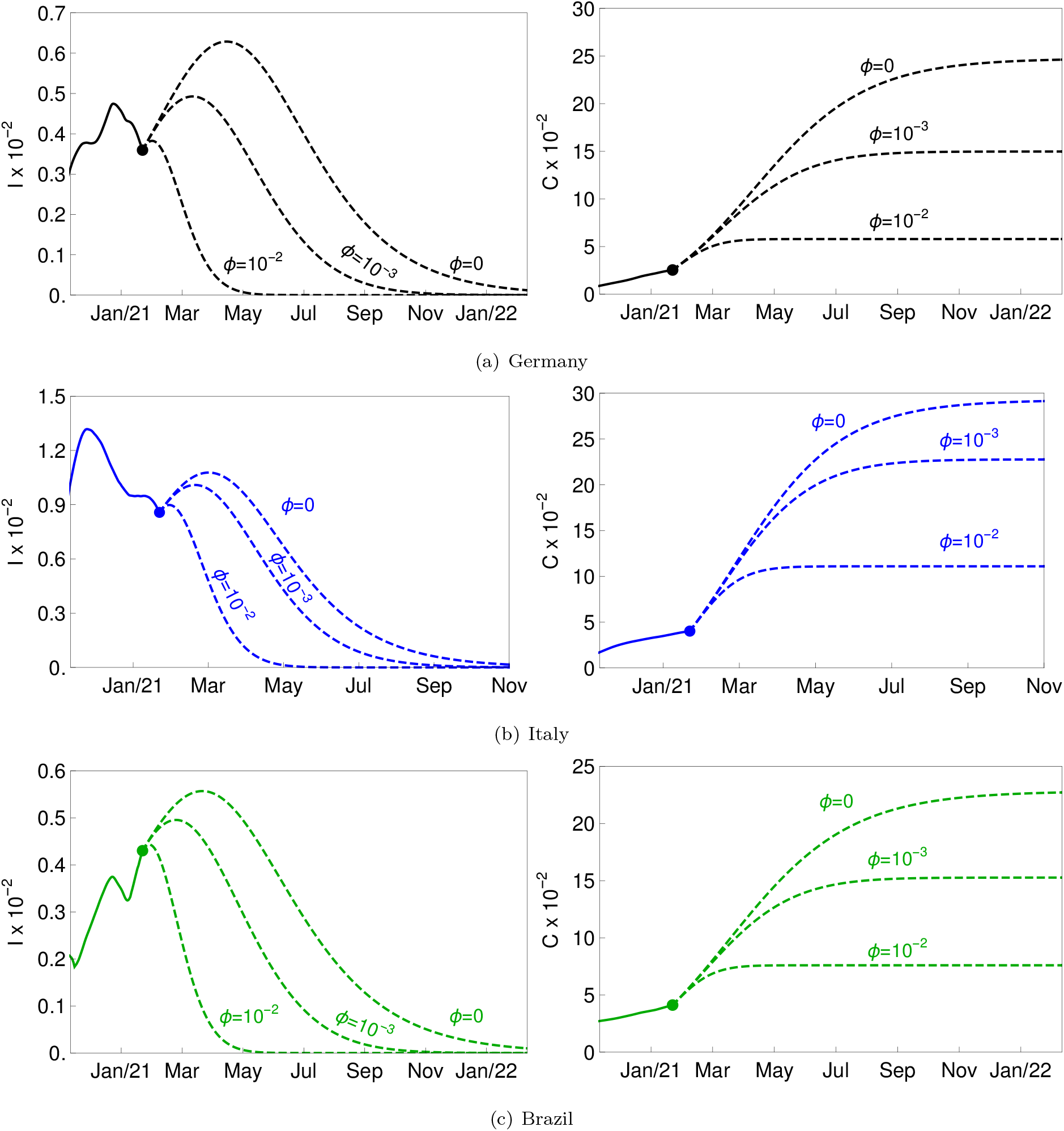
Effect of vaccination in Germany (a), Italy (b) and Brazil (c) with *β*_ger_ = 0.227, *β*_ita_ = 0.230 and *β*_bra_ = 0.224, respectively. The continuous lines stand for real data whose final point is marked as a solid point in the each figure. Dashed lines yield numerical simulations from this point on for three different vaccination coefficients, yielding *ϕ* = 0, 10^*−*3^ and 10^*−*2^.

**Figure 20:**
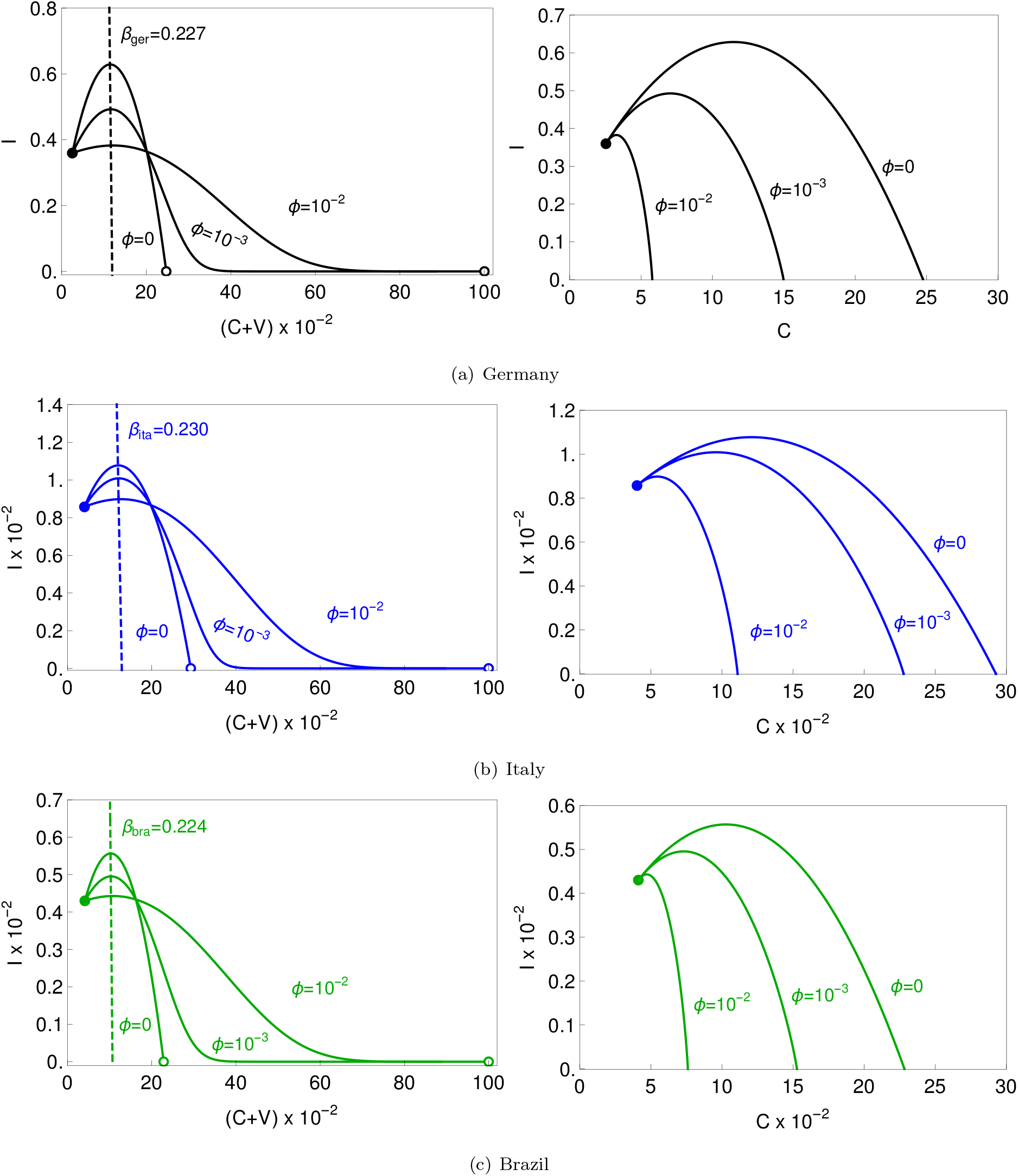
Effect of vaccination employing the maps *I*-(*C* + *V*) and *I*-*C* in Germany (a), Italy (b) and Brazil (c). The dashed line on the *I*-(*C* + *V*) map stands for infectious rate *r* = 1 for *β*_ger_ = 0.227, *β*_ita_ = 0.230 and *β*_bra_ = 0.224.

An interesting point that can be evaluated is the influence of vaccination coefficient *ϕ* on the total infected population *C*(*t* → ∞). Fig. 21 presents this analysis where one can see that the higher number of cases is achieved with the absence of vaccination (*ϕ* = 0). Moreover, the sensitivity of total infected with respect to vaccination rate is given by the curve slope. The higher sensitivity occurs for lower vaccination rates.

**Figure 21:**
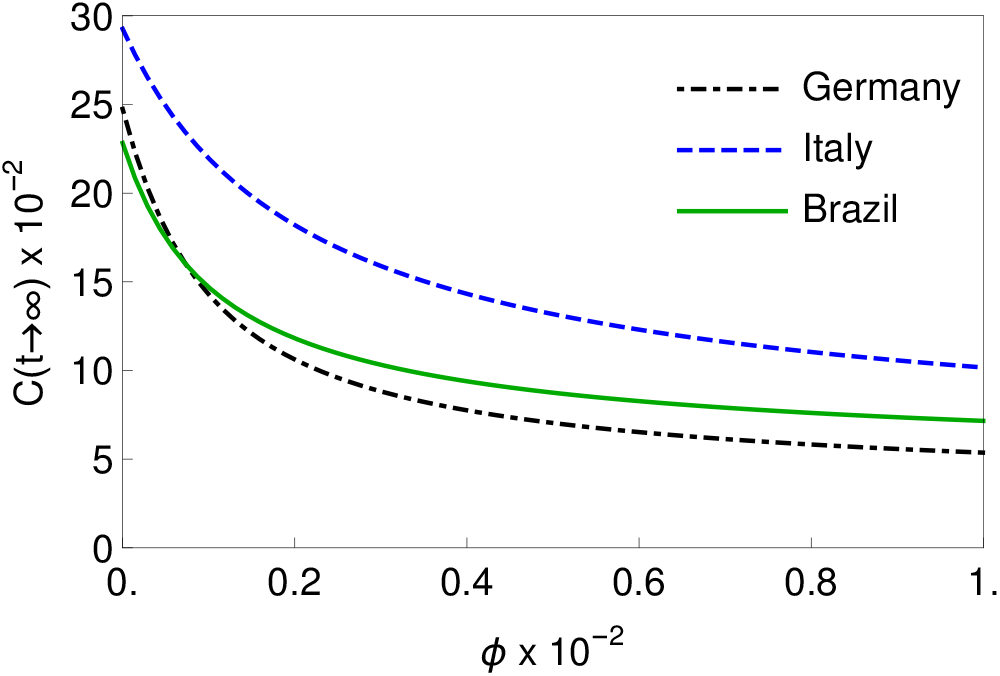
Effect of vaccination rate *υ* on the total cases *C*(*t → ∞*).

The effect of vaccination is summarized in Table 1 showing the total infected population, total deaths and the date when the infected population reaches *I* = 10^*−*4^ for the three vaccination frameworks. The death rate *µ* is adopted based on the average of the final 90 days: 1.86% for Germany, 3.95% for Italy, and 2.68% for Brazil. Without vaccination, the total infected population reaches 24.75%, 29.27% and 22.83% in Germany, Italy and Brazil, respectively. If the vaccination is implemented with 10^*−*2^ rate, the number of total deaths can drop to 35%, 67% and 59% in these three countries, respectively. The estimated date to reach *I* = 10^*−*4^ shows that the vaccination shortened this period in several months. Therefore, the vaccination drastically anticipates the end of a huge crisis.

**Table 1:** Numerical simulation results summarizing the effect of vaccination on total infected cases and total deaths

| Country | $\phi$ | Total Cases $\times 10^{-2}$ | Total Deaths $\times 10^{-2}$ | Date when $I = 10^{-4}$ |
| --- | --- | --- | --- | --- |
| Germany | 0 | 24.75 | 0.22 | Feb/22 |
| | $10^{-3}$ | 14.29 | 0.14 | Sep/21 |
| | $10^{-2}$ | 5.37 | 0.08 | Apr/21 |
| Italy | 0 | 29.27 | 0.25 | Nov/21 |
| | $10^{-3}$ | 21.97 | 0.22 | Aug/21 |
| | $10^{-2}$ | 10.17 | 0.17 | Apr/21 |
| Brazil | 0 | 22.83 | 0.20 | Feb/22 |
| | $10^{-3}$ | 14.70 | 0.16 | Sep/21 |
| | $10^{-2}$ | 7.16 | 0.12 | Apr/21 |

## 6. Conclusions

This paper proposes a dynamical map to describe COVID-19 epidemics based on the classical SEIRV differential model. This novel map describes COVID-19 dynamics from three populations: active infected, cumulative infected and vaccinated. The discrete-time map is advantageous for several reasons: i) it reduces the number of model variables when compared to the continuous SEIRV model; ii) it is described by only three algebraic equations, being easier to be implemented and to perform parameter adjustments; iii) analytical tools can be employed to define useful information such as the infectious rate and the herd immunization point. The herd immunization point is a function of the transmission rate and the infectious average period, being independent of the mean latent period. Model verification compares the map simulations with the classical SEIR model and real data showing a good agreement. Real data from Germany, Italy and Brazil show that the map is capable to capture the main features of the COVID-19 epidemics, including bell shape and plateaus patterns. Nonlinear dynamics perspective shows to be an interesting approach allowing the analysis of the main features of the COVID-19 epidemics and different patterns are reproduced with proper parameter choices. The effect of vaccination is investigated using the novel map and different models to describe the vaccination campaigns. Results show that proper vaccination rate can dramatically reduce the total infected population. The analytical estimation of the herd immunization point allows the evaluation of the end of the pandemic crisis indicating proper vaccination strategy. Based on these results, the novel map can be employed as a useful tool for COVID-19 scenario evaluation, being an easy alternative to be employed.

## Data Availability

All data referred to in the manuscript is based on official information and based on reproducible by numerical simulations.

## 7. Acknowledgements

The authors would like to acknowledge the financial support of the Brazilian Research Agencies CNPq, CAPES and FAPERJ.

